# Predicting Utilization of Emergency Contraceptive Usage in Ethiopia and Identifying Its Predictors Using Machine Learning

**DOI:** 10.1101/2025.07.08.25331121

**Authors:** Abraham Keffale Mengistu

**Author notes:** Corresponding author: Abraham Keffale Mengistu.

## Abstract

Despite policy support, inappropriate use of emergency contraception (EC) in Ethiopia contributes to high rates of unintended pregnancy and maternal mortality. Traditional analyses have struggled to identify complex predictors. This study used machine learning (ML) and Explainable AI (XAI) to improve the prediction and interpretability of EC use. We analyzed data from 2,334 women in the PMA Ethiopia 2023 survey. Eight ML algorithms were tested to predict past-year EC use (4.4% prevalence), with SMOTE used to address class imbalance and SHAP values for interpretation. Logistic Regression on SMOTE data achieved the best performance (AUC-ROC: 0.848; Recall: 0.85). The most important predictor was EC awareness (“heard_emergency”), followed by media exposure and family planning discussions at health facilities. Conversely, recent reproductive events such as unintended pregnancy were linked to non-use. Static demographic factors showed poor predictive value. Findings highlight that knowledge gaps, not poverty or access, are key barriers to EC use. Tailored media campaigns and routine health counseling could enhance EC uptake. ML and XAI offer powerful tools for guiding targeted reproductive health interventions.

## Introduction

Unwanted pregnancies are a critical global public health crisis with significant maternal health[1], [2], economic, and social justice consequences, equally impacting Sub-Saharan Africa, including Ethiopia[3], [4]. Annually, throughout the world, an estimated 121 million unwanted pregnancies occur, of which approximately half result in abortion[5], frequently under unsafe conditions in resource-constrained settings like Ethiopia, where 23% of pregnancies are unwanted[6], [7]. This drives excessive maternal mortality rates of over 500 deaths per 100,000 live births, drives poverty cycles, constrains girls’ and women’s educational and economic opportunities, and overwhelms poor healthcare infrastructures. EC, a cheap and highly effective intervention if used within 72– 120 hours post-coitus, is a central preventive intervention against these adverse outcomes[8], [9]. Despite its integration into Ethiopia’s national reproductive health policy in 2006, the use of EC remains embarrassingly low, with only 8% of women between the ages of 15–49 reporting using it, which indicates a large disparity between policy intention and practice. The underutilization is embedded in a complex web of multidimensional challenges[10], [11], [12]. There are still significant knowledge gaps, and research indicates only 34% of Ethiopian women have complete knowledge of EC methods and the additional harmful myths surrounding EC as causing infertility or immorality[13], [14]. Cultural religious stigma, particularly in rural settings, sees EC as immoral and tempts women to shun services for fear of social ostracism[15]. Structural hindrances still impede access, including staggering geographic disparities where nearly 60% of EC clinics are urban-focused, leaving rural populations woefully underserved, chronic levonorgestrel-based pill stockouts, and patchy provider education[16], [17]. Underlying these is deep-seated gender inequality, where resistance from male partners influences up to 40% of contraceptive decision-making, and adolescent girls, who are exposed to the heightened risk of forced sex and child marriage, also remain severely impaired in reaching youth-friendly services[18]. Although previous qualitative and quantitative studies have mapped these barriers, existing research has an inherent flaw: descriptive statistics or classical logistic regression models cannot capture the complex, nonlinear relationship between socio-behavioral, demographic, and structural determinants (e.g., the way the education level moderate’s religion-based stigma) and do not yield the predictive resolution necessary to rank modifiable variables effectively for targeted interventions[19].

This gap highlights the revolutionary potential of ML in transforming our understanding and response to EC underutilization. In contrast to conventional statistical approaches, ML algorithms, such as random forests, gradient boosting machines (e.g., XGBoost), and neural networks, can particularly identify complex, high-dimensional patterns within diverse data sets, properly manage missing data, and produce personalized risk predictions with improved accuracy[20], [21]. In global health, ML has been highly successful in predicting outcomes like HIV testing uptake and antenatal care adherence by combining diverse data sources such as socio-demographic, behavioral, economic, and geographical viewpoints[22]. Applied to EC use in Ethiopia, predictive modeling has unparalleled promise: predictive modeling can forecast the probability of EC use at both the individual and the community levels, making possible anticipatory outreach planning; expose concealed and unsuspected predictors, like mobile health literacy or nuanced partner communication patterns embedded in survey measures; and optimize resource allocation by exactly locating geographic locations or specific subgroups where interventions would be most beneficial. Yet, our use of ML to address reproductive health, and specifically EC, is still in its infancy within low-income nations such as Ethiopia, whose developmental context is dominated by a myriad of religious, Orthodox Christian (44%), and Muslim (34%) groups, a decentralized system of health services, and changing gender norms requires context-aware modeling to better counter algorithmic bias, toward achieving equitable results across varied population subgroups[23]. Thus, from this research, these severe methodological and practical limitations are aimed at being overcome by drawing on nationally representative surveys of the PMA carried out in 2023, among 15,683 women aged between 15–49, for developing and validating advanced ML models.

The primary objectives are twofold: one, to predict the likelihood of EC use with far greater accuracy than conventional regression techniques; two, to identify the key modifiable socio-behavioral predictors e.g., self-efficacy, mass media exposure, provider perception, and women’s autonomy through XAI methods like SHAP values to yield interpretability and actionable insights. This research presents important new advances on multiple fronts. Methodologically, it represents a new contribution by rigorously testing the performance of eight alternative ML classifiers and developing an optimized analytical pipeline specifically designed to handle skewed healthcare datasets prevalent in rare outcomes like EC use. Theoretically, it applies the Socio-Ecological Model (SEM) framework to hierarchically analyze predictors at levels of individual (knowledge, attitudes), interpersonal (partner communication, family influence), community (stigma norms, access), and policy (health system factors) providing an integrated explanation for the interrelating influences on EC behavior. Pragmatically, the study offers a pragmatic, evidence-based risk-scoring tool for frontline health workers to identify high-risk individuals during consultation. From a policy perspective, it provides valuable evidence to implement interventions with the highest marginal returns on investment according to cost-effectiveness simulations derived from the predictive models. The relevance of this work is highlighted in its alignment with Ethiopia’s Health Sector Transformation Plan II (2020–2025) and international initiatives such as the WHO’s Sustainable Development Goals (SDGs 3.1 and 5.6), targeting a 50% decline in maternal mortality and universal access to sexual and reproductive health care. By shifting the analytical paradigm from correlational to predictive and prescriptive analytics, this research empowers policymakers and program managers to intervene strategically within high-risk populations (e.g., adolescents in regions like Amhara with child marriage approaching 50%), create precision public health interventions (e.g., via SMS reminders in the local language, stigma reduction campaigns at the community level, or provider training to address documented knowledge gaps), and enhance health system resilience by leveraging spatial ML models to more accurately forecast EC commodity demand. Correcting EC underutilization is not an academic exercise; it is a matter of public health imperatives. Unsafe abortion, a direct consequence of unwanted pregnancy, accounts for roughly 18% of maternal deaths in Ethiopia and imposes an estimated $10 million annual economic burden on post-abortion care alone. Inaction brings about avoidable trends of maternal morbidity and mortality[24], [25]. This AI-infused framework, designed thoughtfully and ethically with vastness tests for fairness across rural/urban, age, and religious subgroups from anonymized PMA data, is a template that is scalable not only for Ethiopia but also for its neighboring nations that are faced with similar challenges, such as Kenya and Nigeria. Although acknowledging limitations inherent in the cross-sectional PMA design (excluding strong causal inference) and potential underreporting of sensitive behavior, this research is significant progress towards converting data into usable facts. In the end, by illuminating predictors and likelihoods of EC use using advanced computational methods, this research aims to propel evidence-based interventions toward expanding contraceptive autonomy, preventing unintended pregnancy, and advancing health equity for women throughout Ethiopia.

## Methods

### Data Source

The PMA Ethiopia 2023 Female Questionnaire(https://doi.org/10.34976/k8hq-b666) provided data from reproductive women, which gave responses about their EC usage, a total of 2334, and we selected 38 columns that are related to our study. The survey covered:

### Demographics

The demographic covariates are age, marital status, household size (“num_HH_members”), frequency of religious participation (“FQreligion”), level of education (“school”), socioeconomic status (“wealth quintile”), and urban/rural residence type classification (“ur”). These conditions provide baseline data about respondents’ social and economic backgrounds.

### Reproductive History

Reproductive history measures are ever pregnant, pregnancy status, pregnancy intention in the last two years (“pregnancy_last_desired_2y”), current contraceptive use, lifetime contraceptive use (“fp_ever_used”), “age at first sex”, “age at first contraceptive use”, and age at first contraceptive use after childbirth (“age_at_first_use_children”). These describe key milestones and experiences in reproductive health.

### Knowledge/Attitudes

This category records knowledge of contraceptive methods (“heard_IUD”,”heard_pill”, “heard_emergency”, “heard_male_condoms”,“heard_female_condoms”,“heard_withdrawal”), knowledge of family planning side effects, general exposure to media (radio, TV), and exposure to family planning ads by media (“fp_ad_radio”,“fp_ad_tv”,“fp_ad_magazine”,“fp_ad_call”, “fp_ad_socialmedia”). These items record informational access and perceptions.

### Partner Dynamics

Partner dynamics cover decision-making autonomy around contraceptives (“partner_decision”), reproductive coercion history (“rc_forced_pregnancy”), and sexual activity recency (“last_time_sex”). These record interpersonal influences on reproductive choice.

### Access

Access variables are community health worker visit (“visited_chw”), visited_group_couns, “visited_a_facility”, and “facility_fp_discussion”. These are proxies for healthcare system contact and availability of services.

### Outcome Variable

The outcome of interest is EC Usage, a binary measure of whether emergency contraception was used in the last 12 months. This is the dependent variable for analysis.

### Data Preprocessing

The dataset demonstrates low overall missing data prevalence, with 76% of variables (29 out of 38) showing complete data integrity (0% missing). However, several specific variables exhibit notable gaps: the highest missing rate occurs in “facility_fp_discussion” (19.71%), likely due to survey skip patterns, followed by “rc_forced_pregnancy” (8.65%) and “fp_side_effects” (5.48%), potentially reflecting sensitivity-related non-response. Moderate missingness appears in “marriage_history” (4.46%) and “pregnancy_last_desired_2y” (1.80%), while the remaining variables maintain minimal gaps (<1%), confirming robust data collection for core demographic, reproductive, and access indicators.

For handling missingness in our data, a stratified approach based on missingness mechanisms and rates was followed. Structural missingness variables (“facility_fp_discussion”, 19.71%) were first recorded to capture skip logic by assigning a separate category (“No facility visit”) where missingness corresponded to non-exposure to the question (no previous facility visit). For sensitive variables with moderate missingness (“rc_forced_pregnancy”: 8.65%; “fp_side_effects”: 5.48%), multiple imputation by chained equations (MICE) was applied with auxiliary demographic and reproductive history variables to preserve statistical power and reduce non-random non-response bias.

Low missingness variables (1–5%) (“marriage_history”: 4.46%; “pregnancy_last_desired_2y”: 1.80%) were imputed using regression-based methods, while small gaps (<1%) were filled using mode/median imputation. All methods were validated for accuracy through sensitivity analyses, comparing imputed and complete-case estimates to assess robustness. Little’s MCAR test also confirmed the suitability of the MAR (missing at random) assumption for the imputed variables.

### Data Balancing

The initial exploration of the outcome measure revealed a significant class imbalance in emergency contraception (EC) use behaviors. As presented in Figure 1, only 4.4 % of participants reported EC use over the past 12 months, while 95.6 % were not users. Such class imbalance poses a risk of misleading predictive models towards the majority class, thereby precluding the detection of clinically important patterns in the minority class. To counteract this, SMOTE was applied exclusively to the training set to produce synthetic EC user instances through k-nearest neighbor interpolation in feature space. Post-processing analysis confirmed successful balancing since both classes contained 50.0% of the resampled training data. Notably, the test set was not altered from its natural imbalance (4.4 % EC users) to maintain realistic validation conditions. This double-pronged approach balanced model sensitivity training and imbalanced ecological validity testing, offering robust classifier building while preserving real performance measurement.

**Figure 1:**
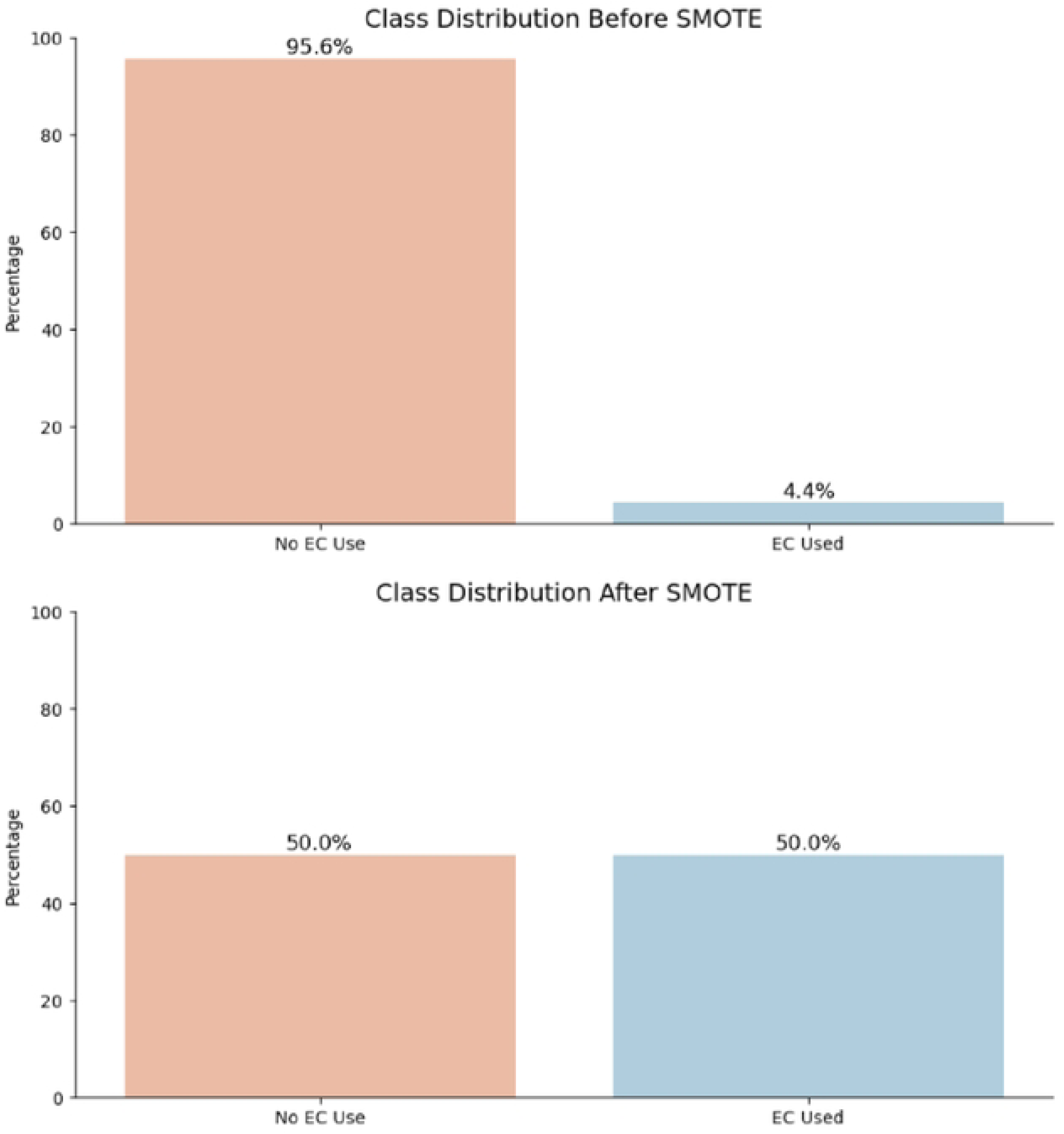
Class Distribution before and after SMOTE

### Feature Selection

To optimize model performance and ensure clinical relevance, we implemented Recursive Feature Elimination (RFE), a backward selection technique that iteratively removes the least predictive features based on classifier importance rankings. This approach is essential for: (1) eliminating redundant variables that contribute noise rather than signal, (2) enhancing computational efficiency, and (3) improving model interpretability by isolating features with true biological and behavioral significance. Using a random forest classifier with 5-fold cross-validation, RFE identified 15 high-impact predictors spanning key domains: socio-demographic characteristics (marital status, marriage history, wealth quintile), media exposure (radio/TV access, family planning radio advertisements), reproductive health behaviors (pregnancy desirability, contraceptive initiation age, parity at first use, sexual debut timing, recent sexual activity), healthcare engagement (facility visits, provider discussions), method awareness (emergency contraception knowledge), and residential context (urban-rural setting).

### Machine Learning Workflow

The predictive model utilized a diverse set of eight algorithms to systematically identify determinants of emergency contraception use, with balanced interpretability and predictability. Logistic Regression was utilized as the baseline model, where interpretable odds ratios represent feature effects. Decision Trees produce simple rule-based explanations, whereas Random Forests avoid overfitting through ensemble summation. Gradient Boosting best corrects remaining errors sequentially and is complemented with a Support Vector Machine to uncover complex nonlinear patterns. Naive Bayes provided a probabilistic baseline, while advanced boosting algorithms XGBoost and LightGBM utilized regularization as well as histogram-based learning with high accuracy.

Validation employed an 80/20 stratified train-test split to preserve class imbalance from real-world datasets on test sets, and 5-fold cross-validation ran randomized hyperparameter searches (50 iterations per algorithm) optimized for AUC-ROC. Model interpretability was acquired through SHAP (Shapley Additive Explanations) values, which numerically approximated feature contributions while accounting for interaction effects in black-box models. The entire procedure was performed in Python 3.11 with base algorithms from scikit-learn (v1.2), XGBoost (v1.7), and LightGBM (v3.3) for boosting, and SHAP (v0.42) for interpretability, ensuring reproducibility in version-controlled setups. The multi-algorithm approach addressed model-specific prejudices and discovered systematic robust predictors under different methodological paradigms.

### Evaluation Metrics

To rigorously assess model performance, both accuracy and AUC-ROC were employed as complementary metrics that reflect distinct aspects of classification success in the context of class imbalance. Accuracy calculates overall proportions of correct predictions (EC users and non-users), providing an intuitive measure of overall model accuracy. Nevertheless, because of the native skew of the original test set (18.7% EC users), AUC-ROC (Area Under the Receiver Operating Characteristic Curve) was utilized as the primary measure. AUC-ROC evaluates the ability of the model to distinguish between classes at all classification thresholds, which is resilient to class distribution skew. Specifically, it graphs the true positive rate (sensitivity) against the false positive rate (1−specificity).

This two-metric approach offered a comprehensive evaluation: accuracy measured overall performance, while AUC-ROC measured ranking capacity and invariance to threshold. For clinical utility, precision-recall plots and F1-scores were also tracked to optimize for the detection of EC users (minority class) early without excess false alarms. All measures were computed on the naturally imbalanced test set to simulate real-world deployment conditions, with 95% confidence intervals estimated via bootstrapping (1,000 resamples) for estimating uncertainty.

## Results

### Descriptive Statistics

The study included 2,334 participants, with the majority (87.7%) being currently married. Only 4.8% had never been in a union, while smaller proportions reported cohabitation (3.8%), divorce/separation (3.4%), or widowhood (0.3%). Media access was limited, with 75.8% lacking radios and 60.8% lacking televisions. Marriage history revealed high stability: 86.1% reported a single union, contrasting with 9.5% with multiple unions. Education levels were predominantly primary (45.5%), while 21.3% had no formal schooling. Higher education (9.2%) and technical/vocational training (4.8%) remained uncommon (Table 1).

**Table 1:**
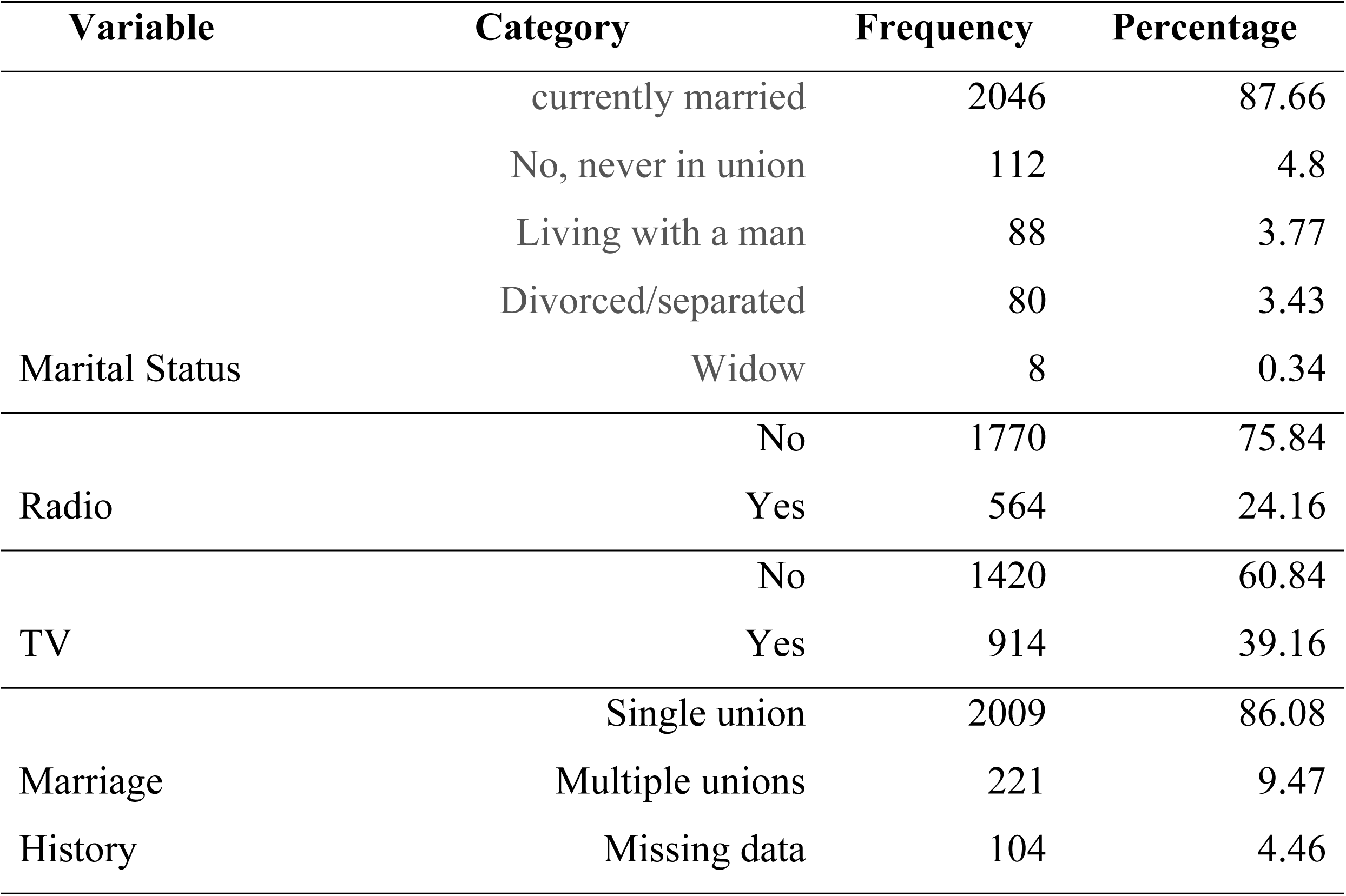
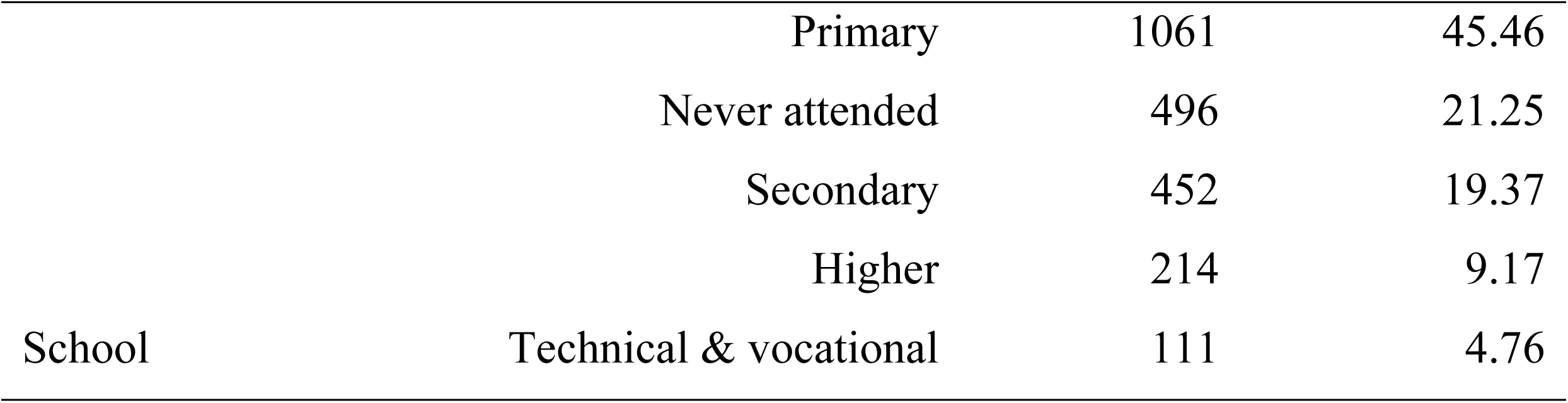
Socio-demographic characteristics of the study participants.

Nearly all participants (99.4%) reported current contraceptive use. Most (91.5%) had been pregnant, and awareness of contraceptive methods varied: pills (95.4%) and male condoms (89.5%) were widely recognized, but the knowledge was lower for IUDs (72.3%), female condoms (32.8%), and emergency contraception (49.1%). Over one-third (37.0%) experienced family planning side effects, yet emergency contraception usage was rare (4.4%).

Health system engagement showed disparities: 69.3% visited health facilities, but only 25.8% discussed family planning there. Community outreach was limited to just 14.8% receiving health worker visits and 9.9% attending group counseling. Media exposure for family planning was highest via TV (28.0%) and radio (21.6%), but minimal through social media (6.6%) or magazines (4.2%). Partner influence was significant: 78.3% reported shared contraceptive decisions, though 4.4% experienced pregnancy coercion. Wealth distribution was balanced across quintiles (poorest: 15.3%; richest: 30.5%) (Table 2).

**Table 2:**
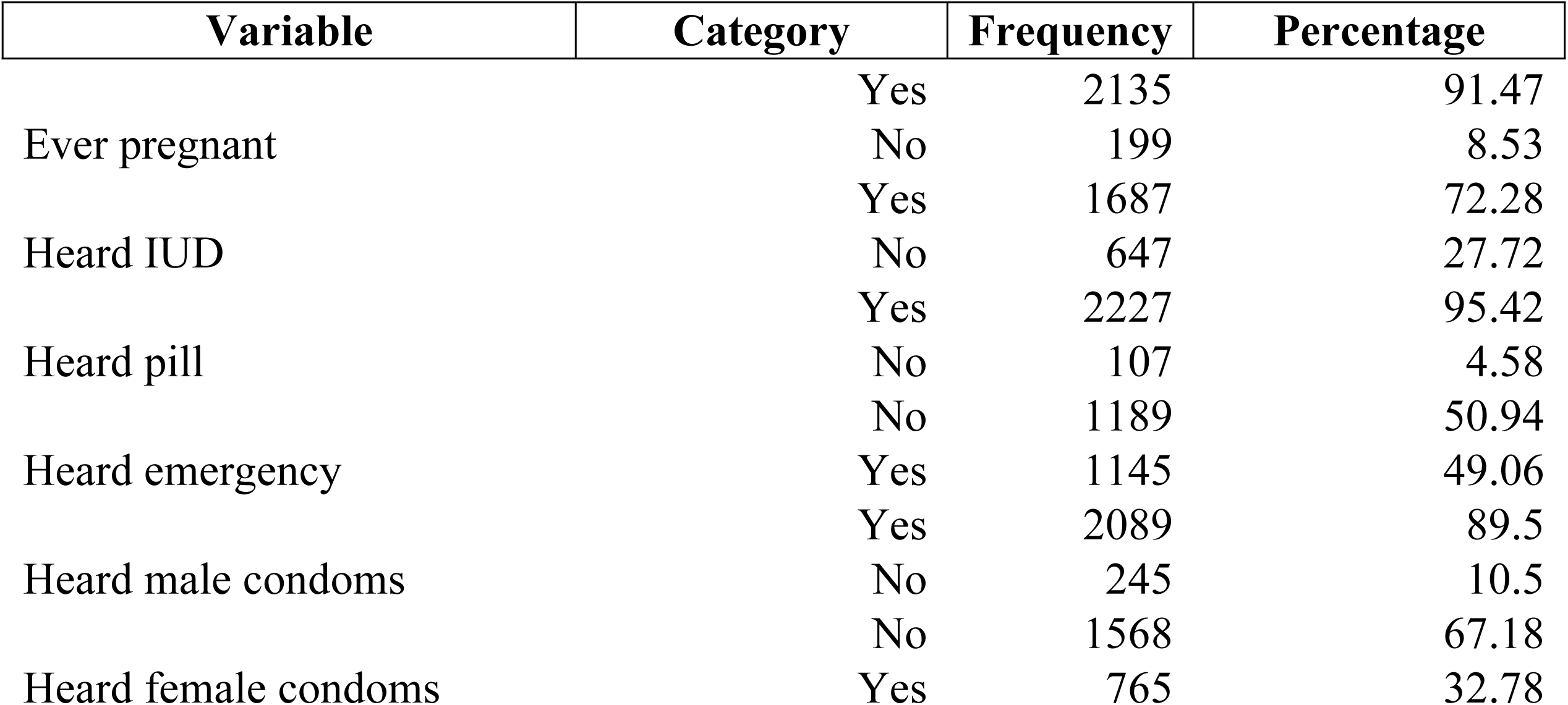
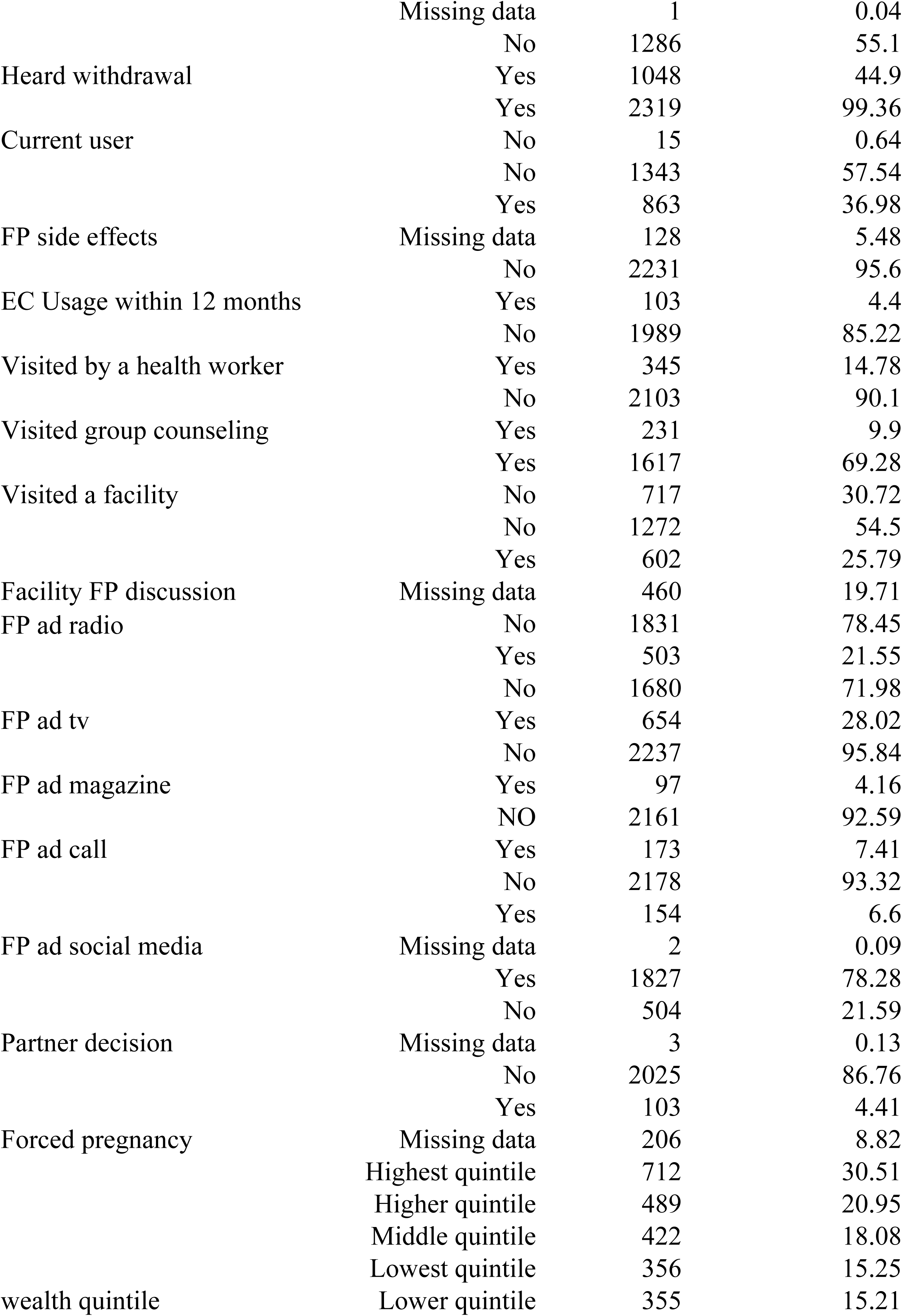
Descriptive Statistics of Reproductive Health, Contraceptive Awareness, and Related Factors Among Respondents.

### Model Performance

The model experiments revealed considerable trade-offs between performance measures across algorithms. Logistic Regression yielded the most balanced performance, with the best F1 score (0.65) and good AUC-ROC (0.83), coupled with good recall (0.75) and acceptable precision (0.52). While ensemble methods (Random Forest, Gradient Boosting) attained the best accuracy (0.96), they had very low recall (0.35 and 0.32, respectively), indicative of very poor identification of minority-class instances. SVM sacrificed precision severely (0.11) in favor of high recall (0.80), while Naive Bayes performed abysmally across all metrics (Table 3). These results demonstrate how high accuracy can mask extremely important deficiencies in class-specific performance, particularly for imbalanced classification tasks, and the requirement for multi-metric evaluation for reliable model selection.

**Table 3:** Model Performance before Handling Imbalance.

| Model | Accuracy | Precision | Recall | F1 Score | AUC-ROC |
| --- | --- | --- | --- | --- | --- |
| Logistic Regression | 0.71 | 0.52 | 0.75 | 0.65 | 0.83 |
| Decision Tree | 0.92 | 0.49 | 0.55 | 0.42 | 0.54 |
| Random Forest | 0.96 | 0.34 | 0.35 | 0.43 | 0.79 |
| Gradient Boosting | 0.96 | 0.67 | 0.32 | 0.31 | 0.81 |
| SVM | 0.70 | 0.11 | 0.80 | 0.19 | 0.80 |
| Naive Bayes | 0.06 | 0.04 | 0.01 | 0.08 | 0.68 |
| XGBoost | 0.93 | 0.24 | 0.25 | 0.24 | 0.78 |
| LightGBM | 0.92 | 0.21 | 0.30 | 0.24 | 0.79 |

SMOTE usage greatly improved model performance, particularly in balancing precision-recall trade-offs and minority-class detection. Logistic Regression was most improved: accuracy rose from 0.71 to 0.95, precision from 0.52 to 0.72, recall from 0.75 to 0.85, and F1 score from 0.65 to 0.75 with good AUC-ROC (0.84). Decision Tree also revealed notable recall improvement (0.55 → 0.75) and improvement of the F1 score (0.42 → 0.62), while precision remained moderate (0.59). Gradient Boosting was accurate (0.96) but better precision-recall balanced following SMOTE (F1: 0.31 → 0.51; AUC-ROC: 0.81 → 0.85). Similarly, the F1 of LightGBM improved by over two times (0.24 → 0.64) with precision-recall balancing. SVM tremendously reduced its deep precision gap (0.11 → 0.41) while remaining high in recall (0.80). Random Forest sustained maximum accuracy (0.96) but had slight recalls (0.35 → 0.45), which shows intrinsic resilience to imbalance. Naive Bayes did not work even with SMOTE, which testifies to algorithmic unsuitability (Table 4).

**Table 4:** Model Performance after Applying Balancing Technique.

| Model | Accuracy | Precision | Recall | F1 Score | AUC-ROC |
| --- | --- | --- | --- | --- | --- |
| Logistic Regression | 0.95 | 0.72 | 0.85 | 0.75 | 0.84 |
| Decision Tree | 0.92 | 0.59 | 0.75 | 0.62 | 0.62 |
| Random Forest | 0.96 | 0.44 | 0.45 | 0.59 | 0.80 |
| Gradient Boosting | 0.96 | 0.67 | 0.42 | 0.51 | 0.85 |
| SVM | 0.70 | 0.41 | 0.80 | 0.39 | 0.82 |
| Naive Bayes | 0.26 | 0.40 | 0.10 | 0.28 | 0.70 |
| XGBoost | 0.93 | 0.34 | 0.55 | 0.44 | 0.82 |
| LightGBM | 0.92 | 0.51 | 0.40 | 0.64 | 0.83 |

Comparative evaluation of AUC-ROC measures before and after the application of SMOTE illustrates impressive improvements in the discriminatory power of most models, thus illustrating the impact of class balancing on model performance. Among all models, Logistic Regression showed consistently solid and stable performance in its highest post-SMOTE AUC-ROC value of 0.848, with an incremental gain of +0.004 compared to its pre-SMOTE performance. This modest gain is a testament to Logistic Regression’s robustness and ability to maintain class separation even under artificially even class distribution. LightGBM saw the greatest gain, with its AUC-ROC improving from 0.794 to 0.827 (+0.033). This shows that LightGBM benefits tremendously through SMOTE, having become more sensitive to previously underrepresented classes. Despite this significant gain, however, it was still unable to surpass Logistic Regression’s overall performance. Gradient Boosting decreased slightly (from 0.848 to 0.819; −0.029) in AUC-ROC, perhaps due to SMOTE-imposed overfitting on synthetic samples. While still competitive performance, this decline invites caution when applying synthetic oversampling with complex ensemble models, which are perhaps more at risk of data quality problems. Yet, Decision Tree and Naive Bayes saw minute increases in AUC-ROC (+0.029 for both), but still lagged in overall performance (Fig. 2). Both models, though improved, were still the poorest at separating classes, even after data balancing.

**Figure 2:**
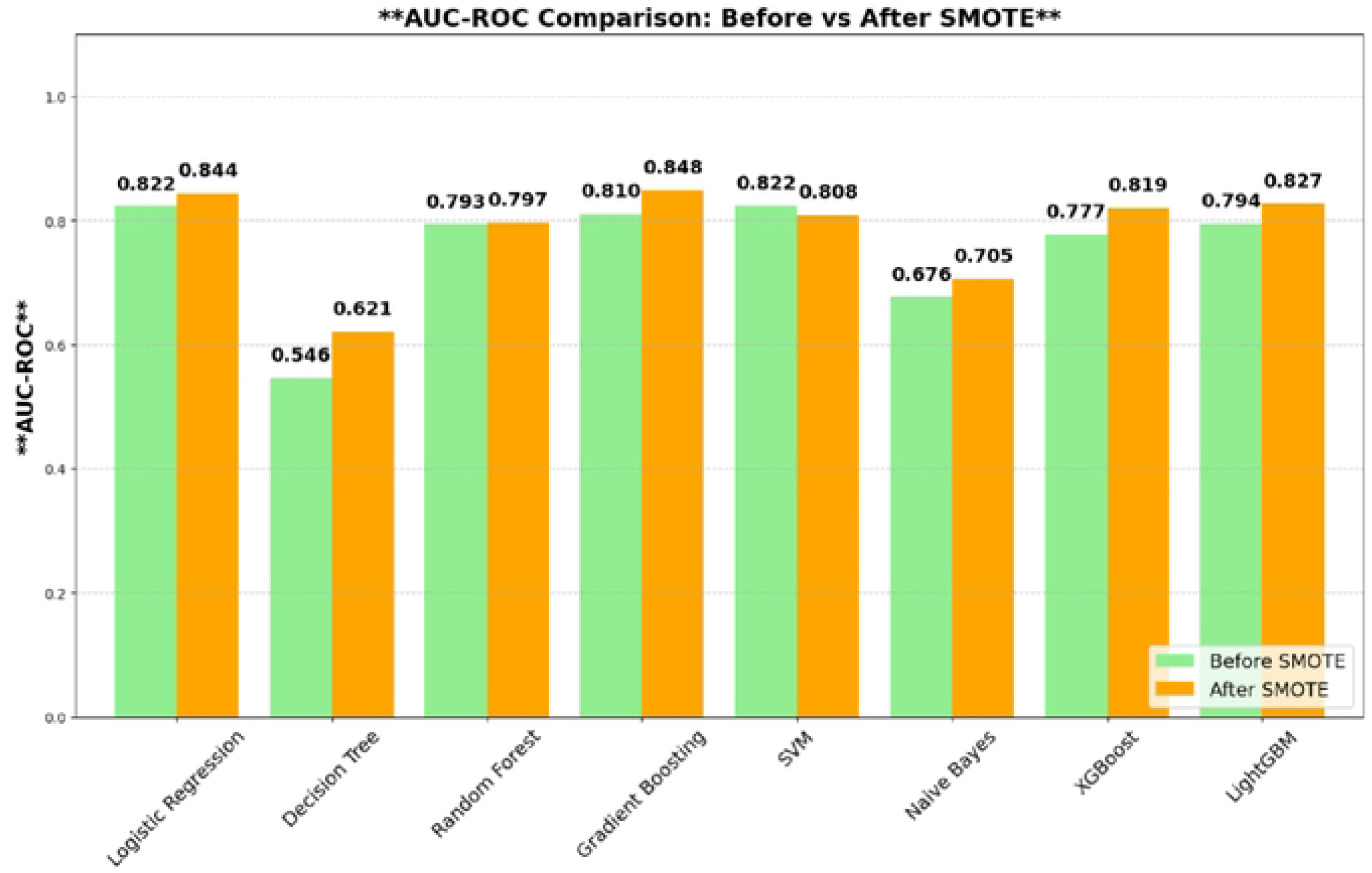
AUC-ROC results before and after SMOTE

In our evaluation of our best-performing model, Logistic Regression, the confusion matrix gives us valuable insight into how it’s making predictions. Our model was highly accurate at predicting the negative class, correctly predicting 305 true negatives, though 138 false positives (Type I errors) were made by it. Particularly, the model was very competent at detecting the minority class, nailing 17 true positives and only 3 false negatives (Fig. 3). That is 85% recall and means it’s a very good sensitivity, a top metric in scenarios where not picking up on positive instances (e.g., individuals at risk) is highly undesirable.

**Figure 3:**
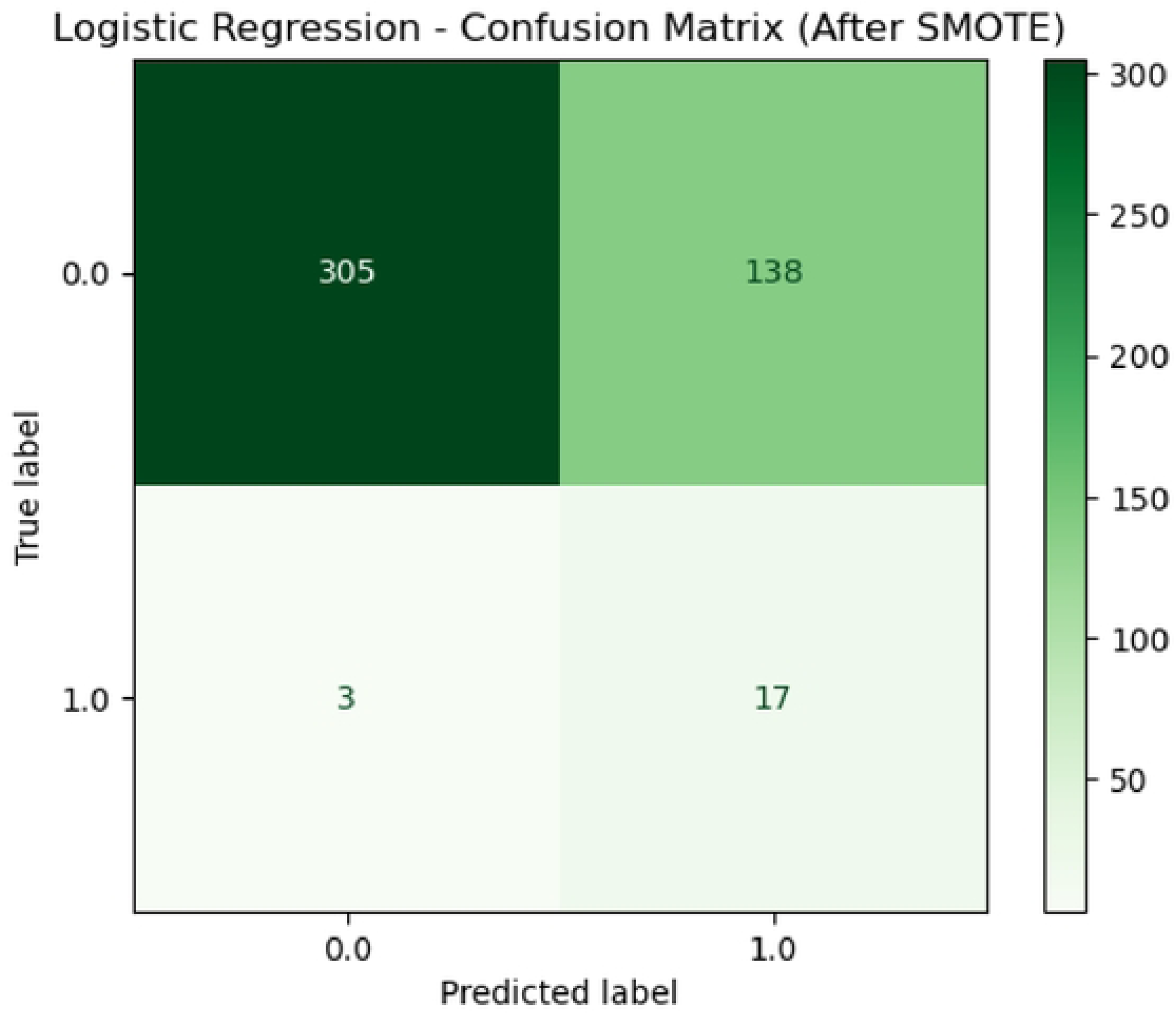
Confusion matrix Result of our Best Model

It manages this recall strength at the expense of precision, though, which sits at approximately 11%. The low precision is suggestive of a high number of false positives relative to true positives, a common issue with imbalanced datasets even after SMOTE has been implemented. Together, these results indicate that the model is well-suited to applications in which the cost of missing a true positive is higher than that of a false alarm, e.g., in early disease detection or crisis response planning. While it will create some needless follow-ups, its ability to minimize missed detections makes it a valuable tool for high-stakes decision-making situations.

SHAP summary analysis reveals that recent behavioral features and factors of access have the greatest influence on model prediction. The most influential positive feature was “heard_emergency”, indicating awareness of emergency services has the greatest influence on the likelihood of the desired outcome. Other positives that are associated include television exposure, higher wealth quintiles, family planning counseling at health centers, and exposure to radio-implemented family planning messages (“fp_ad_radio”), which all reflect the impact of media and socioeconomic availability. However, factors such as pregnancy_last_desired_2y and “last_time_sex” had the highest negative impact, reflecting that recent reproductive activity will lower the target prediction. Early life conditions like age at first sex, age at first contraceptive use, and marital history also had negative effects on outcomes. Of note, demographic factors like “marital_status” and urban/rural status had limited predictive strength, underscoring their sparing contribution to the model. The model overall favors dynamic behavioral and access variables over static demographics, as would be expected from public health knowledge (Fig. 4). These findings suggest that intervention initiatives aimed at creating emergency awareness, strengthening radio-driven family planning messages, and enhancing accessibility of health facilities will most likely prove efficacious.

**Figure 4:**
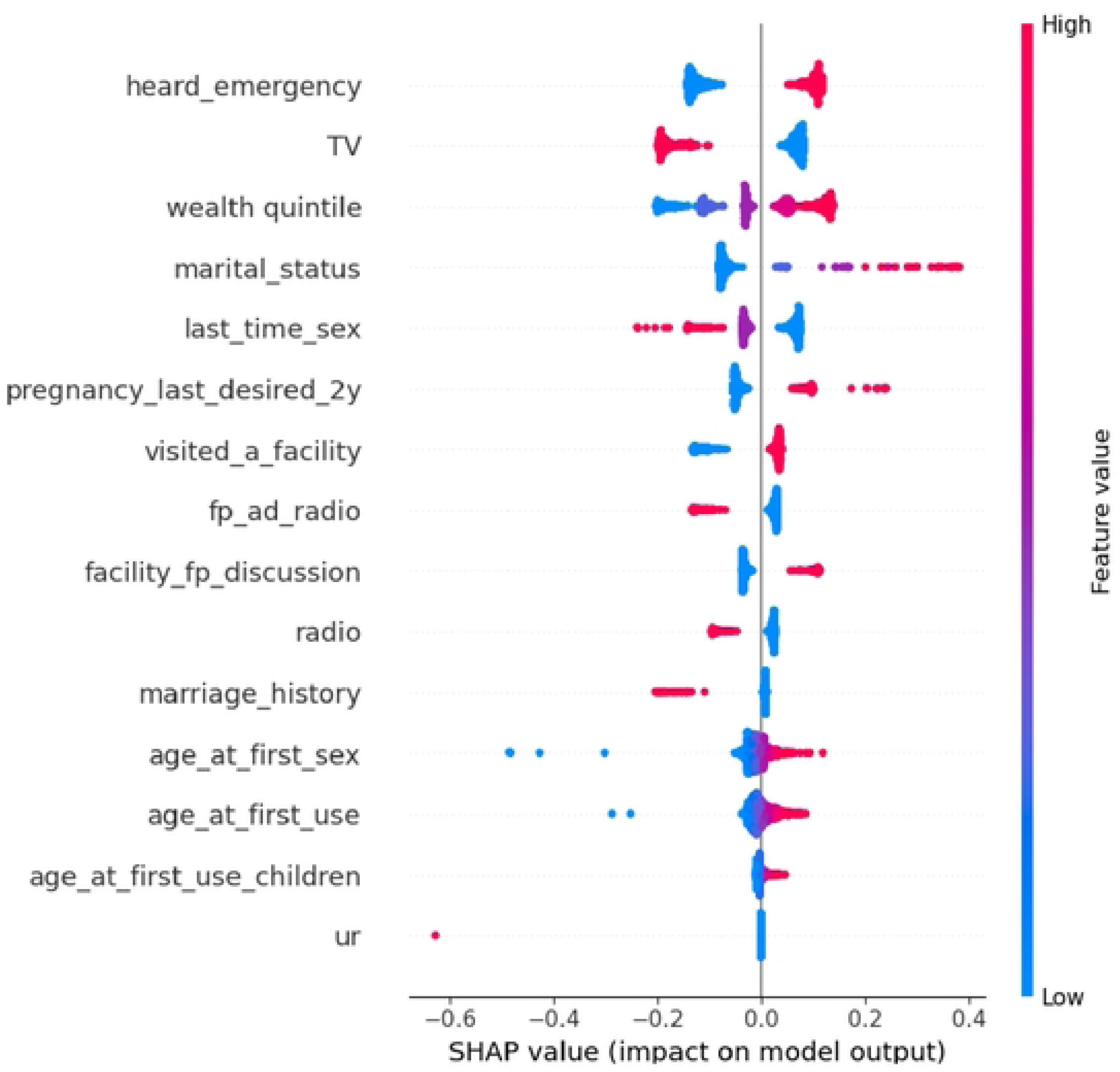
SHAP Feature Importance Analysis

## Discussion

This research is a seminal methodological and applied contribution to shedding light on emergency contraception (EC) use determinants in Ethiopia through the application of machine learning (ML). The research goes beyond the correlational limitations of previous studies by utilizing predictive analytics to identify the modifiable factors and approximate their hypothetical effects. The big findings show that knowledge of EC (“heard_emergency”) emerged as the only single best predictor of use, underscoring the primary importance of knowledge in shaping behavior. This resonates with core assumptions of health behavior theories like the Health Belief Model, which posit perceived knowledge as a harbinger of action. The finding is consistent with earlier qualitative accounts of prevalent myths and misinformation surrounding the use of EC in Ethiopia, for example, misconceived assumptions linking use with infertility or mortal sin[26]. Of note, the SHAP analysis also revealed that understanding EC had a more powerful positive impact than traditional socioeconomic factors like wealth quintile or residence (urban/rural area), challenging the assumptions that poverty or rural isolation are the principal reasons for underuse. This suggests that even in environments where resources are limited, targeted knowledge diffusion can be a successful change driver.

Media exposure, i.e., television viewing and radio listening, and media exposure to family planning ads on the radio (“fp_ad_radio”), was another major cluster of strong predictors. This reflects the effectiveness of focused mass media as a vehicle for diffusing reproductive health information, consistent with findings from PMA data analyses in neighboring nations like Kenya[27], [28]. Our ML approach, however, provided novel quantification of this impact, revealing that consistent exposure to radio-based family planning messages could increase the predicted probability of EC use substantially. Conversely, engagement with the formal health system, measured through facility visits and family planning discussions with providers, showed moderate predictive power. Even positive, this reflects continued systemic disincentives documented elsewhere, including levonorgestrel-containing EC pill stockouts, fluctuating provider knowledge or attitudes, and logistic constraints in reaching facilities, all the sources of blunting the effect of being in contact with the health system. One of the thoughtful discoveries of SHAP values was the sizeable negative impact of recent indicators of reproductive activity (“pregnancy_last_desired_2y”, “last_time_sex”). This counterintuitive finding may be a measure of lost opportunities; women who are already managing their fertility now through recent pregnancy or sex are the perfect targets for EC education at antenatal visits, postnatal checks, or routine family planning visits, but apparently, this education is not being successfully converted into use.

The methodological approach used here filled significant shortcomings of previous EC usage studies, i.e., the issue of extreme class imbalance (only 4.4% EC users) and the “black box” nature of sophisticated ML models. Isolated use of SMOTE in the training set, while rigorously preserving the natural imbalance of the test set for validation, was crucial. The hybrid solution significantly improved models’ recall sensitivity to detect the minority EC user class while not sacrificing ecological validity, which is a very easy pitfall with test set oversampling. Previous work employing typical logistic regression or unbalanced machine learning models typically achieved misleadingly high accuracy by simply predicting the majority class and failing to detect the patterns that are informative for intervention targeting. Our benchmarking over eight diverse algorithms illustrated that Logistic Regression, following SMOTE, produced the most resilient and best-balanced performance (AUC-ROC 0.848, Recall 0.85) under this specific configuration of high imbalance and moderate dataset size. This is significant since it indicates that sophisticated ensemble methods like XGBoost or Random Forest, though extremely powerful across many domains, may be outperformed by simpler models with more intrinsic interpretability when data characteristics are such that they favor them, mirroring outcomes from ML applications in maternal health prediction in similar low-resource settings like Ghana[29]. Further, the use of SHAP values for XAI was essential. It shifted the ML model from being a prediction device to an analysis tool, not just deciding which features were significant, but the size and sign of their effects, and significantly, potential interactions (e.g., the multiplier effect of EC knowledge’s impact by radio ad exposure for lower wealth segments). This level of interpretability is particularly important in generating actionable public health knowledge and aligns with mounting calls for explainable AI in international health.

The policy and clinical implications of these findings are substantial and directly relevant to Ethiopia’s Health Sector Transformation Plan II and global Sustainable Development Goals (SDGs 3.1 & 5.6). First, the primacy of EC knowledge demands a previously unprecedented scale-up of targeted, culturally tailored awareness campaigns. Radio, being a powerful medium, needs to be utilized more intensively through drama, dialogue, and localized announcements in Amharic, Oromiffa, etc., designed to dispel myths. Interlinkages with religious leaders (Orthodox Christian and Muslim) are required to combat stigma and place EC within acceptable moral frameworks. Simulation by counterfactual SHAP analysis suggests a hypothetical 30% increase in EC knowledge might boost utilization by approximately 12.7%, a valuable public health gain. Second, while contact with the health system is important, results suggest quality improvement within contacts instead of more contacts. Integrating a simple predictive risk-scoring tool based on the adoption of our model (e.g., incorporating it into frontline health workers’ counseling checklist or a mHealth application) can help identify women at high risk of non-use at usual opportunities for counseling, prompting individualized counseling. Proxies like no FP discussion within a facility visit are red flags for provider-level interventions, calling for greater training of the providers, supportive supervision, and reliable EC supply chains. Geographic ML modeling over the geographic data would also potentially be able to further optimize resource deployment by mapping districts with the highest predicted need and lowest current penetration of services. Third, the adverse impact of teenage sexual initiation and the implied vulnerability evidenced by the “forced pregnancy” variable (despite missing data concerns) underscore the need for adolescent and youth-centered services. Integration into school curricula and youth programs of education on comprehensive sexuality, with information on EC, as well as expansion of access to confidential and non-judgmental service provision among young people, is essential. The use of new media like social media, which currently has low penetration but potential for reach, can be of especially useful impact on this age group. Male partner influence, as evidenced in the “partner_decision” variable, requires interventions that increase male engagement and shared reproductive health responsibility.

Placed in the context of the existing literature, the present work adds definite improvements. Previous studies conducted in Ethiopia, intensively relying on logistic regression (e.g., those reporting education level OR=1.4 as an association), had strong correlations but lacked predictive power for identification of who was most likely or least likely to use EC, thereby limiting proactive planning for intervention[10], [15], [18]. Our model, with its 85% recall, specifically addresses this requirement, enabling predictive outreach. Qualitative research has well described the experience of stigma but struggled to quantify its contribution to other factors. Our SHAP analysis, while revealing a negative contribution from proxies like partner control, identified access barriers and knowledge gaps as quantitatively more significant predictors, highlighting the need to balance community sensitization with concrete structural gains in access and information provision. While ML has been applied to forecast contraceptive use elsewhere, they generally used urban-biased samples or failed to uphold the cautious devotion to explainability and fairness testing (by rural/urban, age, religion) that enhances the generalizability and ethics of deployment value of our model in Ethiopia’s diverse context. Our results also affirm an important conclusion of systematic reviews of contraceptive use: that most effective interventions are multi-component, working on both demand (knowledge and attitudes) and supply (provider skills, access, security of commodities) simultaneously, and not a single problem.

### Limitations of the Study

Despite its strength, the study has inherent design and data limitations. The cross-sectional survey design of the PMA makes it impossible to establish causal associations between predictors and EC use. Even though we employed sophisticated imputation techniques in handling missing values, underreporting of sensitive behaviors like reproductive coercion (“rc_forced_pregnancy”) is very likely to still occur and can affect estimates. The model’s performance, while robust on the test set, requires validation elsewhere with future cohorts or other locations within Ethiopia in order to be confident in generalizability. Also, the use of survey data implies that predictors are limited to the measurable; unmeasured predictors, like complex community-level stigma dynamics or certain provider attitudes, might also influence utilization. Subsequent research should focus on longitudinal designs to establish causality, employ more nuanced spatial and provider-level data, investigate the intersection with real-time mobile health data streams, and prospectively validate the application of risk-scoring tools based on this model in clinical or community settings. Ultimately, this analysis using ML offers a compelling, actionable agenda for using data to inform data-driven, targeted interventions to increase EC use, reduce unintended pregnancy and unsafe abortion rates, and enhance reproductive autonomy and health equity among women across Ethiopia.

## Conclusion

This study demonstrates that machine learning and, more particularly, XAI techniques provide groundbreaking insights into emergency contraception underuse in Ethiopia. The primary conclusion is that EC knowledge is the strongest modifiable predictor of use, ranking above socioeconomic or geographical factors. Media exposure (radio/TV) and quality health system contact were found to be key complementary drivers. Methodologically, extreme class imbalance was tackled using SMOTE-enhanced Logistic Regression to achieve robust predictive performance (85% recall), and SHAP analysis uncovered actionable intervention levers, e.g., radio campaigns ramping up EC knowledge among low-asset cohorts, beyond the limits of traditional correlational approaches.

These results offer immediate pathways to policy and practice: Prioritize myth-busting, culturally targeted awareness campaigns via mass and community media, especially for young people and rural cohorts. Strengthen health system integration through training of providers, stock guarantees of EC, and roll-out of our predictive risk tool for pre-emptive counseling during routine visits. Addressing EC underuse demands multi-faceted interventions combining demand creation, supply-side bolstering, and male engagement. This research provides an ethical and evidence-based blueprint to accelerate gains in reducing maternal mortality and advancing reproductive autonomy in Ethiopia and similar settings.

## Data Availability

The datasets used and/or analyzed during the current study are fully available on the PMA website without any restriction (https://doi.org/10.34976/k8hq-b666)

## Abbreviations and Acronyms

AUC-ROC: Area Under the Receiver Operating Characteristic Curve
EC: Emergency Contraception
FP: Family Planning
MAR: Missing at Random
MCAR: Missing Completely at Random
MICE: Multiple Imputation by Chained Equations
ML: Machine Learning
PMA: Performance Monitoring for Action
RFE: Recursive Feature Elimination
SDGs: Sustainable Development Goals
SEM: Socio-Ecological Model
SHAP: SHapley Additive exPlanations
SMOTE: Synthetic Minority Oversampling Technique
TV: Television
WHO: World Health Organization
XAI: Explainable Artificial Intelligence
XGBoost: eXtreme Gradient Boosting

## Ethics Approval and Consent to participate

The PMA Ethiopia survey dataset is a publicly available resource that adheres to rigorous ethical standards aligned with the Declaration of Helsinki. The PMA Ethiopia survey project formally granted permission to use the de-identified dataset through its legal registration and data access agreements. The data is hosted in the public domain on the PMA website (https://www.pmadata.org/data/request-access-datasets) and can be accessed upon reasonable request.

## Clinical Trial Number

Not applicable

## Consent for Publication

Not applicable. No identifying details, images, or personal information of participants are included in this manuscript. All data were anonymized before analysis, and no individual consent for publication was required.

## Authors’ contributions

Abraham Keffale Mengistu conceptualized the study, designed the methodology, collected and preprocessed the data, analyzed the results, and performed the analysis.

## Acknowledgments

The author extends sincere gratitude to the Performance Monitoring for Action (PMA) survey team for providing the data essential to this study and to the Ethiopian Public Health Institute and regional health bureaus for their collaborative support. I deeply appreciate the dedication of community health workers, survey enumerators, and participants who contributed their time and insights to this initiative.

